# Outcomes Associated with Rural Emergency Department Provider-to-Provider Telehealth for Sepsis Care: A Multicenter Cohort Study

**DOI:** 10.1101/2022.06.02.22275922

**Authors:** Nicholas M. Mohr, Uche Okoro, Karisa K. Harland, Brian M. Fuller, Kalyn Campbell, Morgan B. Swanson, Cole Wymore, Brett Faine, Anne Zepeski, Edith A. Parker, Luke Mack, Amanda Bell, Katie DeJong, Keith Mueller, Elizabeth Chrischilles, Christopher R. Carpenter, Kelli Wallace, Michael P. Jones, Marcia M. Ward

## Abstract

**Objective:** To test the hypothesis that provider-to-provider tele-emergency department (tele-ED) care is associated with more 28-day hospital-free days and improved Surviving Sepsis Campaign (SSC) guideline adherence in rural emergency departments (EDs).

**Methods:** Multicenter (n=23), propensity-matched, cohort study using medical records of sepsis patients from rural hospitals in a well-established, on-demand, rural video tele-ED network in the upper Midwest between August 2016 and June 2019. The primary outcome was 28-day hospital-free days, with secondary outcomes of 28-day in-hospital mortality and SSC guideline adherence.

**Results:** A total of 1,191 patients were included in the analysis, with tele-ED used for 326 (27%). Tele-ED cases were more likely to be transferred to another hospital (88% vs. 8%, difference 79%, 95% CI 75–83%). After matching and regression adjustment, tele-ED cases did not have more 28-day hospital-free days (difference 0.80 days longer for non-tele-ED, 95% confidence interval [CI] [-0.87]–2.47) or 28-day in-hospital mortality (adjusted odds ratio [aOR] 1.61, 95% CI 0.74-3.57). Adherence with both the SSC 3-hour bundle (aOR 0.80, 95% CI 0.24– 2.70) and complete bundle (aOR 0.81, 95% CI 0.15–4.41) were similar. An *a priori*–defined subgroup analysis of patients treated by advanced practice providers suggested that the mortality was lower in the cohort with tele-ED use (aOR 0.19, 95% CI 0.04–0.90) despite no significant difference in complete SSC bundle adherence (aOR 2.48, 95% CI 0.45–13.76).

**Conclusions:** Rural ED patients treated with provider-to-provider tele-ED care in a mature network appear to have similar clinical outcomes to those treated without.

## INTRODUCTION

### Background

Sepsis is a serious medical condition responsible for over 11 million worldwide deaths annually. It is now the leading cause of in-hospital mortality in the U.S. and comes at a cost of over $62 billion.^1-4^ Early aggressive resuscitation has been shown to improve outcomes in early sepsis, and the *Surviving Sepsis Campaign* (SSC) publishes recommendations to guide early sepsis care.^5, 6^ The Centers for Medicare and Medicaid Services and the National Quality Forum have adapted the SSC guidelines into a nationally reported quality metric called the Severe Sepsis and Septic Shock Management Bundle (SEP-1), and adherence with this bundle of care has been associated with improved sepsis survival.^7-11^

Rural hospitals continue to suffer from sepsis outcome disparities, with low-volume emergency departments (EDs) having 36% higher sepsis mortality than high-volume EDs.^12, 13^ Rural EDs experience staffing challenges, infrequent patients with critical illness, and reduced access to specialists, leading to variable sepsis guideline adherence.^14-18^ Many rural sepsis patients are transferred to other hospitals as part of their care, but even regionalized care does not fully reduce disparities in outcomes.^14, 19-22^

### Importance

One promising strategy to improve rural sepsis outcomes is the use of provider-to-provider ED-based telehealth consultation (tele-ED). Tele-ED care connects a rural clinician to a specialist in a tertiary care center to provide clinical guidance, assist with documentation, arrange interhospital transfer, and support rural EDs caring for critically ill patients.^23^ In small pilot studies, tele-ED has been associated with improved sepsis guideline adherence, possibly attributable both to direct care recommendations and durable training for local medical staff.^24-28^ In addition to tele-ED applications, telehealth has also been used to supplement sepsis care during prehospital care, hospitalization, and post-discharge recovery.^29-33^

### Goals of This Investigation

The objective of this study was to measure the association between tele-ED use in sepsis care and outcomes in a mature, rural tele-ED network. Our hypothesis was that tele-ED care would increase 28-day hospital-free days through the association between tele-ED care and improved SSC guideline adherence. We also hypothesized that we would observe differences in the effect of telehealth by hospital volume and staffing model.

## METHODS

### Study Design and Setting

The <u>TELE</u>medicine as a <u>V</u>irtual Intervention for <u>S</u>epsis Care in <u>E</u>mergency <u>D</u>epartments (TELEVISED) study was a retrospective, multicenter (n=23), propensity-matched comparative effectiveness cohort analysis measuring the association between tele-ED consultation and clinical outcomes in rural ED sepsis patients between August 1, 2016 and June 30, 2019.

Included hospitals participated in a provider-to-provider rural tele-ED network and were in areas defined as rural by the Federal Office of Rural Health Policy.^34^ Nineteen participating centers (83%) were federally designated critical access hospitals (CAHs), and 4 (17%) had an intensive care unit (ICU). The methods of the study have been reported previously (ClinicalTrials.gov NCT04441944), including a detailed description of participating sites.^35^ Initially we planned to include 25 hospitals, but two hospitals were excluded because of a lack of data availability (**Supplemental Figure S1**). This article is reported consistent with the Template for Intervention Description and Replication checklist for Population Health and Policy (TIDieR-PHP), and the study was designed using the Strengthening the Reporting of Observational Studies in the Epidemiology (STROBE) statement.^36, 37^ This project was approved under waiver of informed consent by local institutional review boards.

### Selection of Patients

Patients were identified through a data query of the electronic medical records (EMR) in all participating sites and included all adults (age≥18 years) diagnosed with sepsis during the study period. Sepsis was defined as requiring all of the following 4 criteria: (1) inpatient discharge diagnosis of sepsis based on *International Classification of Diseases, 10*^*th*^ *edition, Clinical Modification* (ICD-10-CM) codes, (2) infection diagnosed in the ED (based on clinical documentation in EMR), (3) organ failure in the ED (defined as a Sequential Organ Failure Assessment Score [SOFA] of 2 or greater or a change of at least 2 in those with chronic disease), and (4) at least 2 systemic inflammatory response syndrome (SIRS) criteria in the ED.^38, 39^ This definition was selected not only to parallel the Sepsis-3 criteria but also to limit the study cohort to those with convincing evidence of infection in the ED who were identifiable with sepsis at the time of the initial ED visit.^40^ Because sepsis diagnosis codes have poor sensitivity, we classified discharge diagnosis codes as sepsis using the Fleischmann-Struzek ICD-10-CM approximation or an explicit sepsis code (R65.20 or R65.21).^41-43^

### Interventions

All sites subscribed to the Avera eCARE hub-and-spoke tele-ED network, based in Sioux Falls, SD. This service provides 24-hour, on-demand, high-definition audio and video consultation with a board-certified emergency physician and experienced ED nurse to over 190 EDs in 13 states, and it is the largest rural tele-ED network in North America. Rural staff may request consultation by activating a button in the ED, and hub staff have access to the local EMR. Standardized nurse-directed triage sepsis screening is done in all participating sites, and hub clinicians use computerized decision-support software to prompt completion of SSC guideline elements. We have reported details of this network previously,^44-47^ and a protocol for standardized sepsis consultation criteria was implemented at all network sites in 2017.^48^

### Measurements

We identified sepsis cases with an EMR query, then trained research assistants abstracted data from both rural hospital and referral hospital (for transferred patients) EMRs using a structured electronic data collection tool in accordance with the methods described by Kaji et al.^49^ Tele-ED consultation was identified by linking EMR records with the tele-ED provider call log, and data abstractors were blinded to the use of tele-ED.

We defined antibiotic appropriateness from independent review by two clinical pharmacists as being consistent with a definition of (1) SSC-defined broad spectrum coverage (primary definition, Gram-positive and Gram-negative coverage consistent with SSC recommendations^50^), (2) source-directed coverage (secondary definition, according to relevant Infectious Diseases Society of America source-specific clinical guidelines for the site of suspected infection^51^), or (3) neither, with disagreements resolved by the study principal investigator. Using this classification, we defined our primary analysis as broad-spectrum coverage, then we conducted a sensitivity analysis with appropriate antibiotics recategorized as adherent if antibiotics met *either* the broad-spectrum or source-directed definitions. We used the ED SOFA score as our severity of illness measure because all elements of the score were available at the time of tele-ED consultation.^38^ Time zero was defined as the first time that SIRS criteria were met in the ED (if not met at the time of ED triage). Patients recognized as having sepsis by hub clinicians were recorded in a sepsis quality improvement database.

### Outcomes

The primary outcome was 28-day hospital-free days, which was calculated as the total number of days within 28 days from the index ED visit that the patient was alive and not in the hospital. This composite measure incorporates both survival and length-of-stay, and any patients who died in the hospital were assigned zero hospital free-days.

Secondary clinical outcomes of interest included 28-day in-hospital mortality, mechanical ventilation use, vasopressor use, 28-day ventilator-free days, 28-day vasopressor-free days, new renal replacement therapy, change in SOFA score over the first 24 hours (delta-SOFA), ED length-of-stay, and time-to-inpatient arrival. Secondary process outcomes included completion of the SSC 3-hour bundle (measure lactate, draw blood cultures before antibiotics, administer broad-spectrum antibiotics, and administer 30 mL/kg crystalloid fluid bolus if lactate is >4 mmol/L or systolic blood pressure is <90 mmHg), completion of the 6-hour bundle (vasopressors for mean arterial pressure < 65 mmHg after fluid resuscitation, repeat lactate if initial lactate > 2 mmol/L), and completion of the entire 3- and 6-hour bundles according to the SSC 2016 guidelines (complete adherence).^50^

Sensitivity analyses were conducted using the 1-hour bundle (3-hour bundle elements completed within 1 hour of diagnosis), broad-spectrum *or* source-directed antibiotics (instead of only allowing broad-spectrum antibiotics to qualify), and a less-restrictive 6-hour time window for all 3-hour and 6-hour bundle elements.

*A priori*–defined subgroup analyses were conducted for (1) patients treated exclusively by local advanced practice providers (APPs) (not including cases treated by rural physicians), (2) patients treated in low-volume hospital EDs (defined as hospitals with fewer than 100 sepsis cases during the study period), (3) patients with SOFA >6, (4) patients whose tele-ED activation occurred early (within 1 hour or 3 hours of sepsis diagnosis, compared with all non-tele-ED cases), and (5) tele-ED patients for whom sepsis was recognized by the hub tele-ED team and included in a quality improvement database (compared with non-tele-ED cases).

### Statistical Analysis

We used a propensity-matched cohort design for the primary analysis, including the following variables presumed to be associated with the use of tele-ED in our propensity score: age, sex, body mass index, past medical history (chronic obstructive pulmonary disease [COPD], cirrhosis, solid organ transplant, cancer, diabetes, dialysis), mode of arrival (private vehicle, ambulance), triage pulse, triage systolic blood pressure, ED SOFA score, initial lactate, suspected source of infection, ED provider type (APP, physician), hospital rurality (4-category Rural-Urban Commuting Area codes^52^), city population, ED annual volume, number of index hospital inpatient beds, presence of an ICU, and distance from the index hospital to the most likely receiving hospital. Because significantly more tele-ED non-exposed patients were available in the analytic data set, we matched tele-ED patients using a variable ratio (nominal ratio 1:2) with non-exposed sepsis patients (using a caliper width of 0.15 to break dissimilar matches) using an optimal matching algorithm to minimize overall Mahalanobis distance. All included patients had outcomes available, but we used multiple imputation (5 data sets) for patients with missing covariates. Our analyses used generalized linear models (GLMs) in our matched cohort, adjusting for variables with persistent imbalances after matching and clustered on pair identifier to account for correlation between matched patients. We used the unmatched cohort for subgroup analyses with small case counts, and sub-analyses were analyzed using GLM regression. We considered p<0.05 as statistically significant using two-sided tests, and a detailed description of our statistical methodology is included in **Supplemental Appendix A**.

#### Sample Size

We powered the study to detect a 10% difference in 28-day hospital-free days, based on a pilot study in the same network in 2016.^25, 53, 54^ Using a paired t-test to reflect the matched study design, we estimated that we needed at least 234 pairs matched 1:1 (as originally planned) between tele-ED-exposed patients and tele-ED non-exposed patients (α=0.05, β=0.20, ρ=0.25, mean=18.5 days, SD 8.2 days). The final analysis used variable matching and conditional regression, so we estimated that our power was adequate if we included at least 234 tele-ED cases. We used all available sepsis cases during the study period, and we conducted all analyses using SAS v. 9.4 (SAS Institute, Cary, NC) or R v. 4.1.2 (R Foundation, Vienna, Austria).

## RESULTS

### Characteristics of Study Subjects

Of 1,191 records included in the analysis, 326 (27%) had tele-ED used during the index ED visit (**Figure 1**). The most common reasons for exclusion were that no infection was recognized in the ED or no organ failure was present in the ED. The median time from ED arrival to meeting sepsis criteria was 3 minutes (interquartile range [IQR] 0–10 minutes). Of tele-ED patients, 266 (82%) had tele-ED consultation within 3 hours after sepsis criteria were met (**Supplemental Figure S2**). The median age was 72 years (interquartile range [IQR] 62–82 years), and 536 (45%) were female. The ED SOFA score ranged from 2 to 13, with a median of 4, and 583 (50%) presented to the rural ED by ambulance. Approximately 57% of total cases came from the 4 highest-volume hospitals in the cohort (**Supplemental Figure S3**). The median 28-day hospital-free days was 23 days (IQR 18–25 days), and 28-day in-hospital mortality was 10% (n=112). Only 11% (n=128) had complete adherence with the 3-hour bundle, and the most common reason for this failure was not receiving appropriate antibiotics within 3 hours (91% of non-adherent cases). Among the 326 tele-ED patients, 44 (13%) were coded as having sepsis cases in the tele-ED hub quality improvement database. For tele-ED patients not identified as having sepsis, the most common case classifications were pneumonia (n=49, 15%) and urinary tract infection (n=29, 9%).

**Fig. 1.**
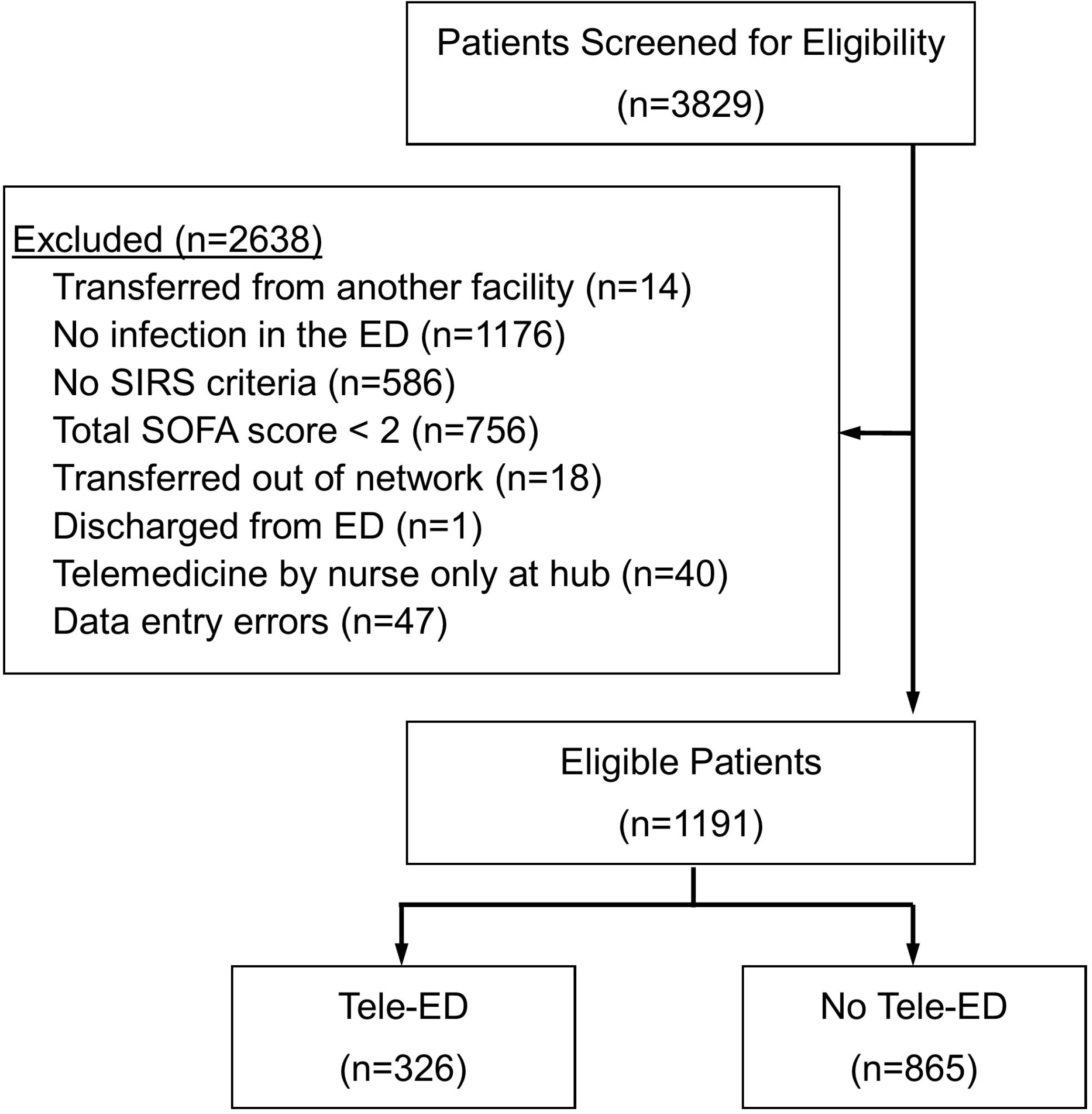

Tele-ED patients were younger, had higher ED SOFA scores and lower systolic blood pressure, and were more likely to have chronic dialysis than non-exposed patients (**Table 1**). Tele-ED patients were more frequently transferred to another hospital (88% vs. 8%, difference 79%, 95% CI 75–83%), and hospital-specific transfer proportion and tele-ED use were strongly associated (**Supplemental Figure S4**).

**Table 1.**
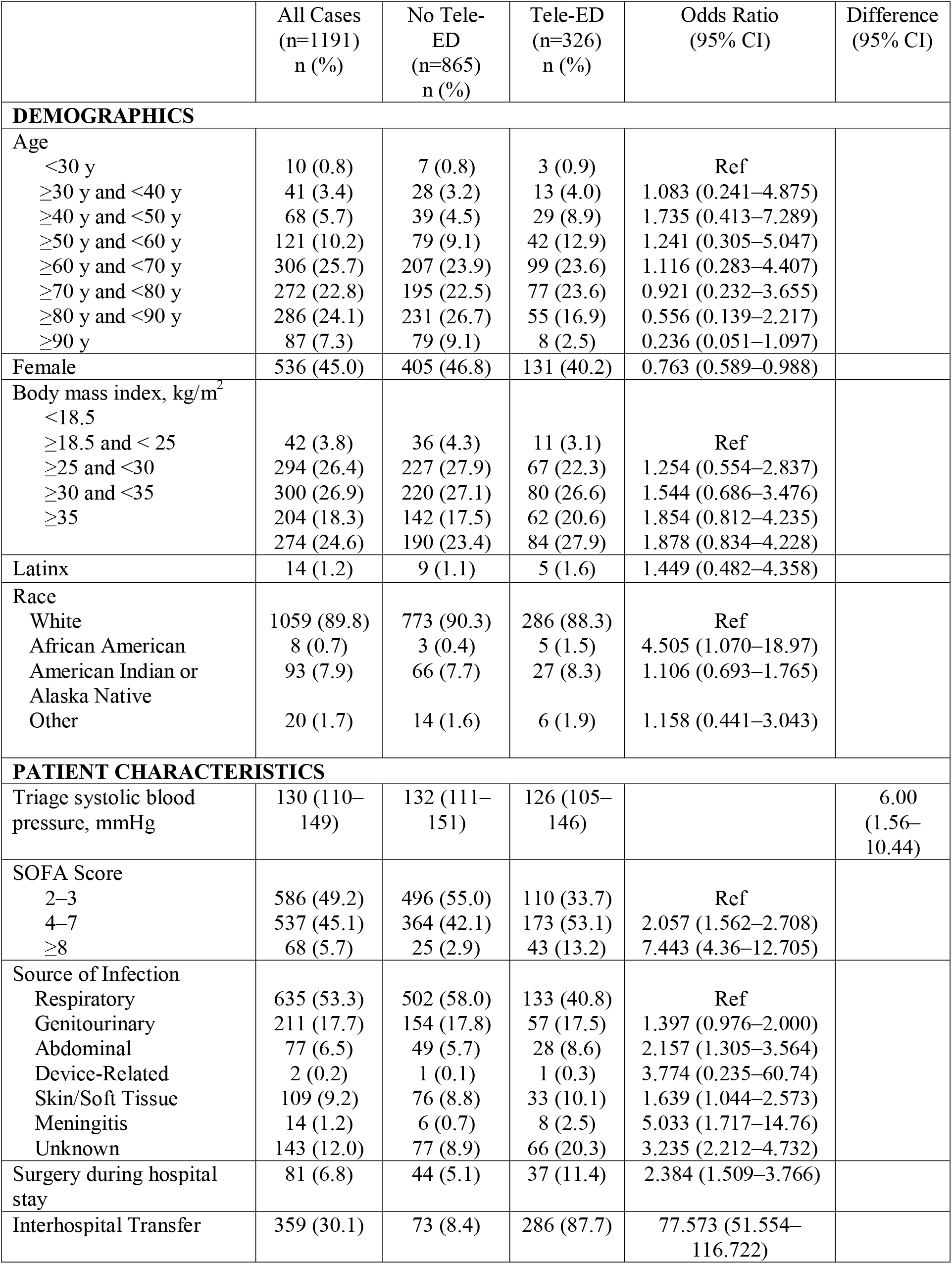
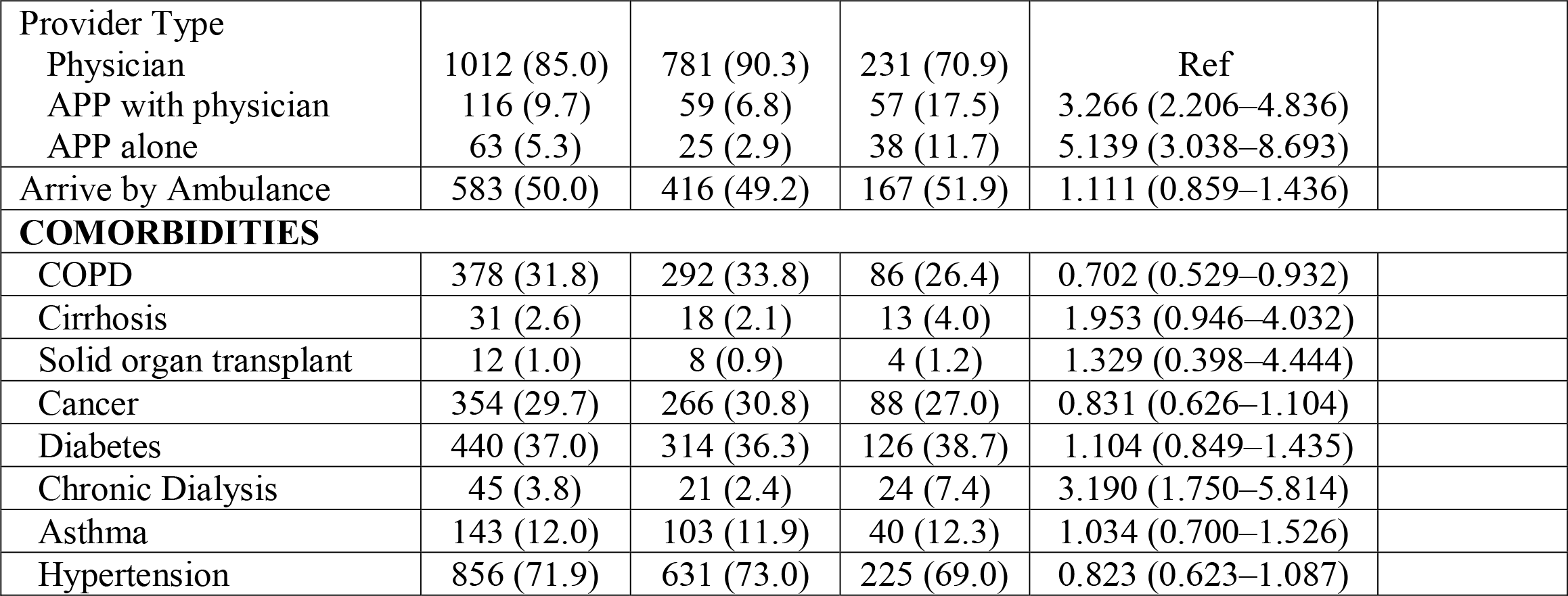
Characteristics of patients.

### Propensity Matching

Propensity scores were higher for tele-ED sepsis patients than for non-tele-ED patients, and the distribution of propensity scores aligned after matching (**Supplemental Figure S5**).

Within the first imputation, we were able to match 242 tele-ED patients with 368 non-tele-ED patients, and standardized mean differences were less than 0.1 for all variables except age, indicating good balance (**Supplemental Figure S6**). Matched patients were had greater severity of illness than the unmatched (**Supplemental Table S1**).

### Main Results

#### Primary Outcome

After propensity matching and regression adjustment, there was no significant difference between tele-ED and non-tele-ED 28-day hospital-free days (difference 0.80 days longer for non-tele-ED, 95% CI [-0.87]–2.47 days, **Supplemental Figure S7**).

#### Secondary Clinical Outcomes

Adjusted mortality for tele-ED patients was not significantly different (adjusted odds ratio [aOR] 0.62, 95% CI 0.28–1.35). There were no significant differences in mechanical ventilation use, vasopressor use, new renal replacement therapy, delta-SOFA score, or 28-day ventilator- or vasopressor-free days. ED length-of-stay was longer in tele-ED patients (median 3.2 vs. 2.5 hours, difference 0.7 hours, 95% CI 0.5–0.9 hours), and tele-ED patients had 1.6 hours longer time-to-inpatient arrival (adjusted median 5.5 vs. 3.9 hours, 95% CI 1.0–2.2 hours), partially attributable to the higher proportion of inter-hospital transfer in tele-ED cases. **Figure 2** shows adjusted and unadjusted clinical outcomes.

**Fig. 2.**
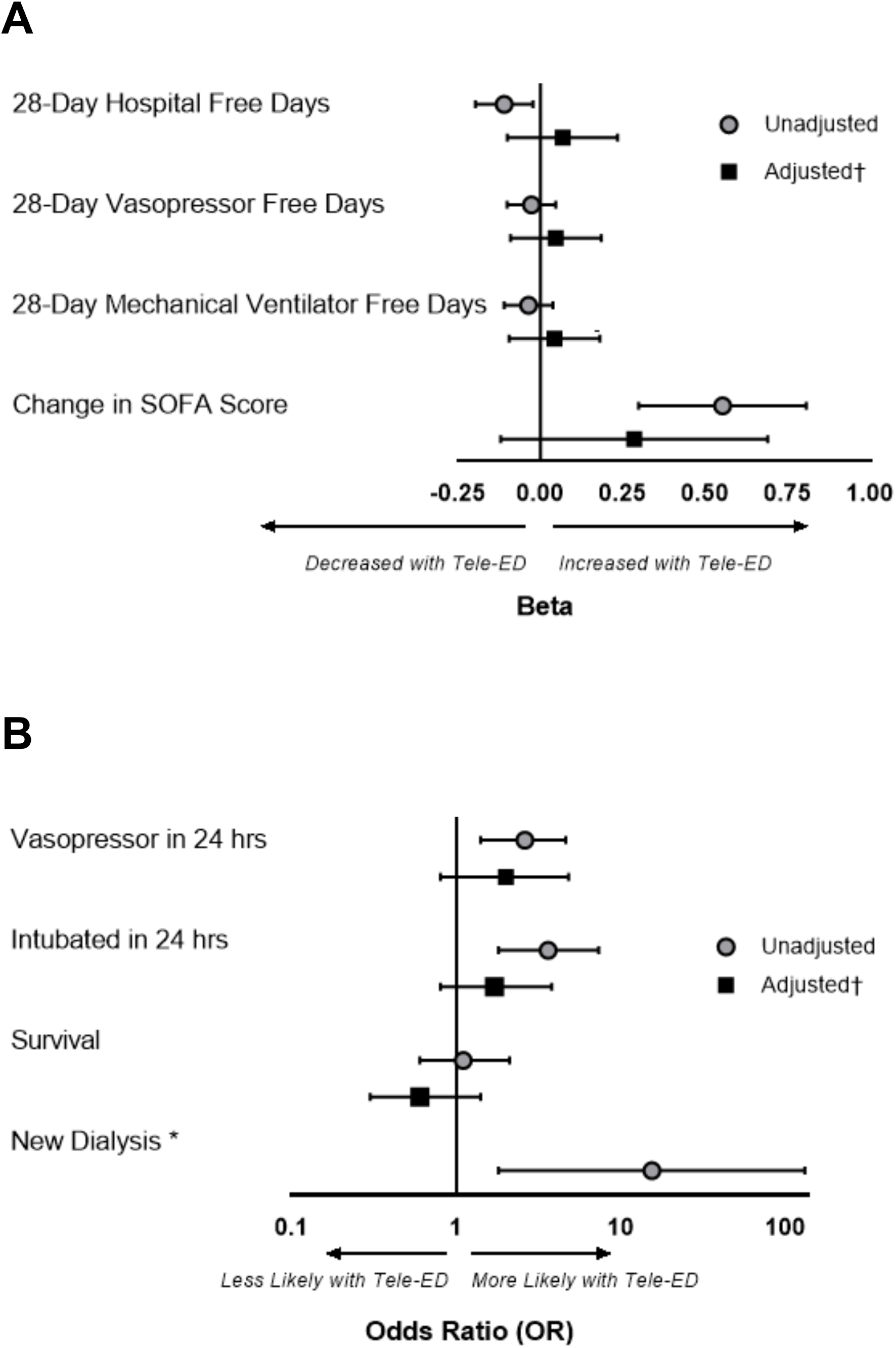

#### Secondary Process Outcomes

n the adjusted model, there was no difference in overall bundle adherence (aOR 0.81, 95% CI 0.15–4.41), 3-hour bundle adherence (aOR 0.80, 95% CI 0.24–2.70), or 6-hour bundle adherence (aOR 0.73, 95% CI 0.33–1.57). Individual bundle element completion is shown in **Figure 3**.

**Fig. 3.**
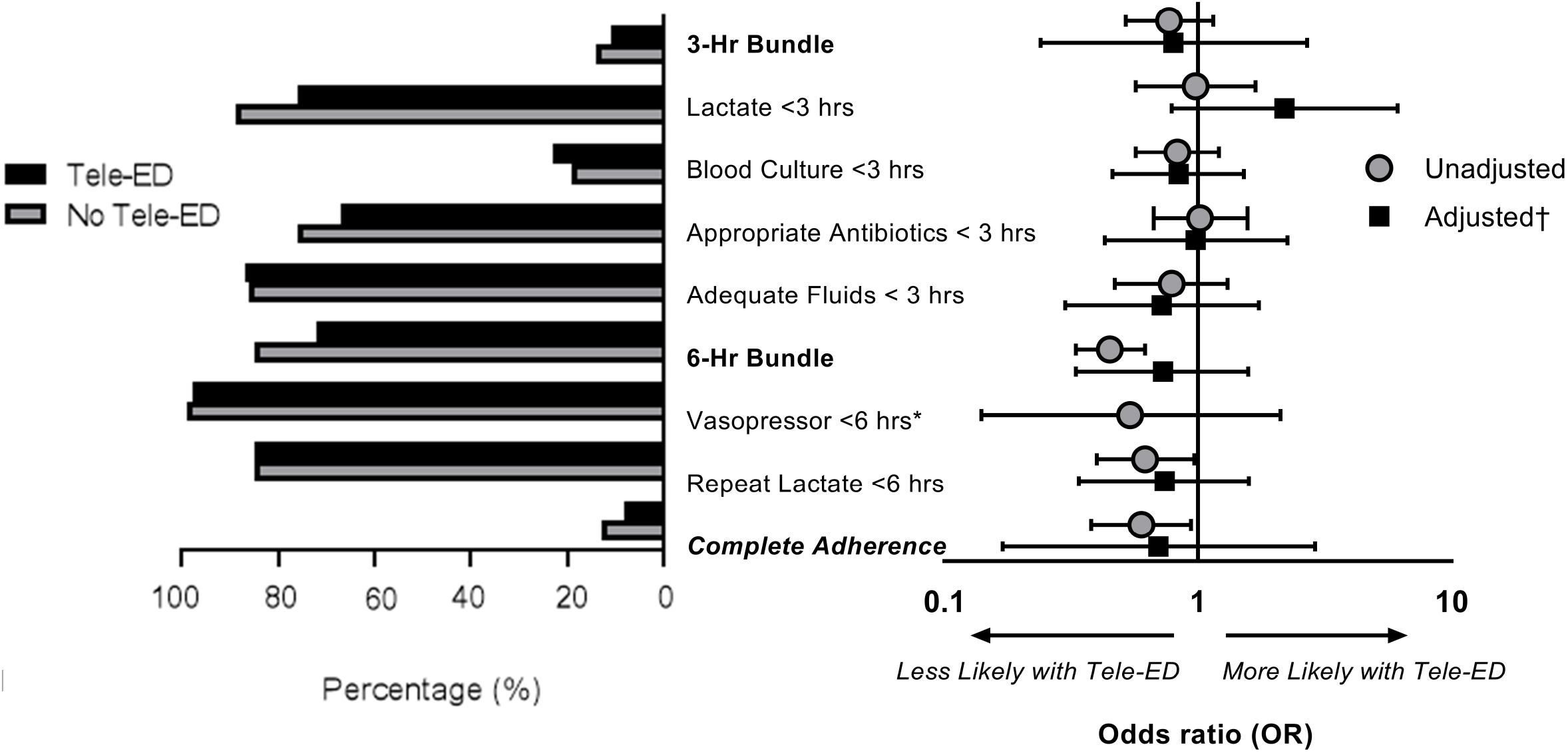

### Sensitivity Analyses

#### One-Hour Bundle

One-hour bundle adherence was uncommon (2.4%), and the probability of 1-hour adherence was not higher for tele-ED patients (OR 0.84, 95% CI 0.36– 1.99). Only 10% of tele-ED patients used tele-ED within 1 hour of ED arrival, however. One-hour adherence was higher in tele-ED cases when tele-ED was consulted within 1 hour (aOR 6.13, 95% CI 1.65–22.72).

#### Antibiotic Appropriateness

Antibiotic adherence increased from 20% to 53% when appropriateness criteria were expanded to include source-directed antibiotics, but the association between tele-ED use and 3-hour bundle adherence remained the same (**Supplemental Figure S8**).

#### Six-Hour Time Window

Broadening the time window for adherence to 6 hours for all 3-hour and 6-hour bundle elements increased the proportion of sepsis patients receiving adherent care by 15%, but tele-ED and bundle adherence was similar (**Supplemental Figure S9**).

### Subgroup Analyses

#### APP Cases

APPs with no onsite physician were much more likely to use tele-ED than physicians alone (60% [APP without physician] vs. 49% [APP with onsite physician] vs. 23% [physician alone]; odds ratio [OR] [APP without physician vs. physician] 5.14, 95% CI 3.04— 8.69; **Supplemental Figure S10**). In fact, 29% of all tele-ED cases were treated by an index ED APP. APPs consulted tele-ED earlier in the ED visit than sepsis patients treated by physicians (1.1 vs. 1.8 hours, difference 0.7 hours, 95% CI 0.4–0.9 hour). Among patients seen by APPs, telemedicine was associated with lower 28-day in-hospital mortality (aOR 0.19, 95% CI, 0.04– 0.90). Complete guideline adherence was not significantly higher in APP tele-ED patients vs. those that did not (aOR 2.48, 95% CI 0.45–13.76, **Figure 4**).

**Fig. 4.**
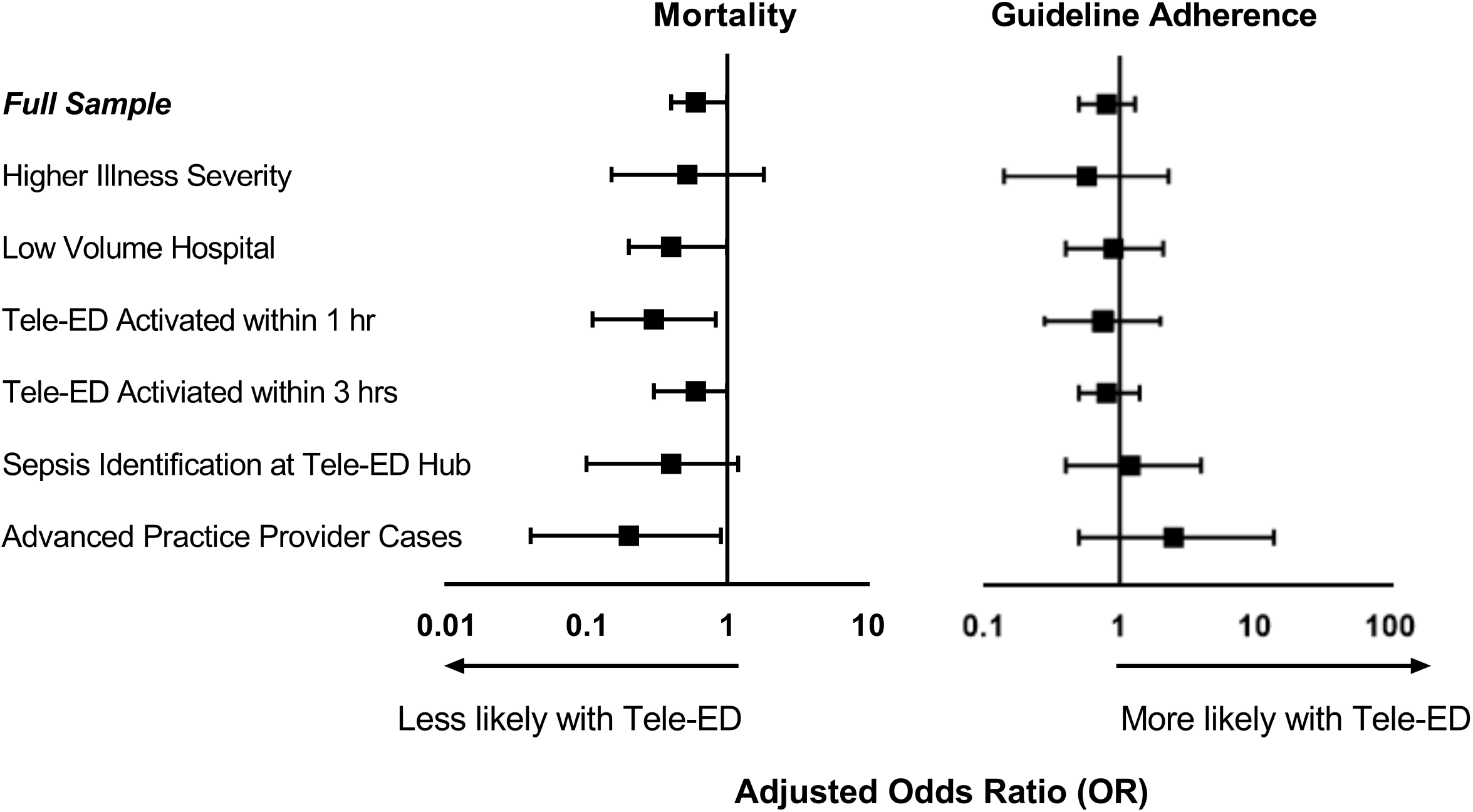

#### Low-Volume Hospitals

Hospitals with low sepsis volumes had similar mortality for tele-ED and non-tele-ED patients (9% vs 13%, relative risk [RR] 0.74, 95% CI 0.42–1.31) and complete guideline adherence (8.6% vs 8.5%, RR 1.02, 95% CI 0.53–1.93). In low-volume hospitals, tele-ED was not associated with significantly lower adjusted mortality compared with non-tele-ED care (aOR 0.44, 95% CI 0.19–1.02) or complete guideline adherence (aOR 0.92, 95% CI 0.41–10, **Figure 4**).

#### Sepsis Patients with Higher Illness Severity

Patients with the highest illness severity (SOFA>6) had higher mortality than those with lower SOFA scores (29% vs. 8%, difference 21%, 95% CI 12–30%). Among those with SOFA>6, however, there was no significant association between tele-ED use and 28-day in-hospital mortality (aOR 0.52, 95% CI 0.15–1.83) or complete guideline adherence (aOR 0.57, 95% CI 0.14–2.29).

#### Early Tele-ED Activation

Sepsis patients who had tele-ED consulted within 3 hours of diagnosis had higher SOFA scores (median 4 vs. 3, difference 1.0, 95% CI 0.6–1.4), lower systolic blood pressure (difference 6.0 mmHg, 95% CI 1.0–10.0 mmHg), and higher transfer proportion (88% vs. 8%, difference 80%, 95% CI 72–87%) than those with later consultation. Replicating the analysis with tele-ED cases being included only if tele-ED was used within 3 hours (versus patients where tele-ED was never used) revealed similar findings (**Figure 4, Supplemental Figure S11, Supplemental Figure S12**). Limiting the tele-ED sample to only those consulted within 1 hour of diagnosis also had similar findings on complete bundle adherence, but lower adjusted mortality (aOR 0.30, 95% CI 0.11–0.83).

#### Sepsis Identification at Tele-ED Hub

Subgroup analysis of only those recorded as having sepsis in the tele-ED hub quality improvement database revealed similar findings (**Figure 4**).

## LIMITATIONS

This study has several limitations. First, while our study was conducted within a national tele-ED network, it remains an observational study. We relied on EMR documentation using variables likely to be recorded accurately, but residual confounding remains possible. Second, all participating sites subscribed to the same tele-ED network with uniform practices and procedures, which may limit generalizability to other networks. Third, the strong relationship between inter-hospital transfer and tele-ED use and the frequency of late tele-ED activation suggests that some tele-ED consultations may have been only for arranging inter-hospital transfer. These consultations could have diluted the effect of tele-ED on clinical or process outcomes, simply because care was delivered without hub recommendations. Fourth, the cohort severity of illness was lower than expected in our power analysis, allowing the possibility that our study was underpowered. We used all available records, however, which resulted in a sample much larger than predicted in our *a priori* power analysis. Fifth, by only focusing on patients who met rigid diagnostic criteria that included identification of infection by the local provider, this study does not address diagnostic error—a significant consideration in sepsis care. This is relevant because improved diagnosis in the tele-ED group, for instance, could introduce bias into the assessment of outcomes. Finally, there has been increasing criticism of the SEP-1 quality metric.^55-57^ We used a clinical outcome as our primary analysis, but SEP-1 elements may not reflect all the ways in which tele-ED hub providers influenced care.

## DISCUSSION

In this multicenter observational cohort study of 1,191 sepsis patients in rural EDs, we did not find that tele-ED as it was used in this network was associated with improved clinical or process-oriented outcomes. This finding was unexpected because prior studies have shown robust improvements in adherence with quality measures when telehealth is used to supplement sepsis care.^24, 25, 27, 58-61^ We also found a strong relationship between inter-hospital transfer and tele-ED use, suggesting that one reason for use of tele-ED may have been shifting the administrative burden of the transfer process to the tele-ED hub. This is especially important because tele-ED consultations often occurred late in the ED stay, which may have reduced the capacity of the tele-ED intervention to influence sepsis treatment. We also observed a hypothesis-generating finding in a subgroup analysis that there may be an association between tele-ED use and sepsis survival in patients treated by APPs, but the small sample in this subgroup and the large effect size draws confidence in this finding into question.

Sepsis is one condition that could be especially sensitive to improvements in care quality from a tele-ED intervention.^62^ Although many rural EDs treat sepsis infrequently, all EDs have the technology required for high-quality early care. Sepsis has a varied presentation, and the permutations of comorbidities, source of infection, and sepsis phenotype could make expert consultation valuable. Finally, sepsis treatment is time-sensitive,^63^ but in contrast to trauma care, appropriate early care is probably more important than timely arrival to a tertiary center.

Prior sepsis tele-ED studies have been smaller-scale demonstration projects early in the development of a telehealth program. Those studies showed that provider-to-provider telehealth was a feasible intervention and that it may be associated with better adherence with standardized care.^24, 25, 27, 58-61^ In a prior study of tele-ED care in trauma, we observed that care changed significantly in hospitals when they adopted tele-ED care, but those changes in care affected both tele-ED and non-tele-ED patients.^44, 64^ In a follow-up qualitative study, rural clinicians cited the secondary education provided by mentored, telehealth-facilitated patient experiences as one of the valuable aspects of the tele-ED service—it changed the way these clinicians practiced.^26^ Tele-ED care stimulates collaboration on standardized protocols, quality improvement, and *ad hoc* conversations with tertiary experts in the tele-ED hub. One reason that tele-ED consultation may not have been associated with improved outcome was that treating clinicians had adopted routine sepsis care customarily recommended by hub providers.

Guideline adherence in this cohort of rural sepsis patients was remarkably low, primarily because of low frequency of administration of timely and appropriate broad-spectrum antibiotics. Sepsis patients presenting with vague symptoms continue to be challenging to diagnose,^65^ and some authors have suggested that patients with atypical sepsis symptoms suffer a greater burden of sepsis care delays.^66^ In our study, tele-ED intervention was activated largely after sepsis recognition, limiting the role that tele-ED could play in the earliest phase of care. Antibiotics may be important even for patients with low illness severity,^67^ but we have shown that the transfer process itself may be associated with delays in appropriate therapy because patients are placed in ambulances for transfer prior to early care being complete.^14^ Few sepsis patients have ongoing resuscitation continued by emergency medicine services, representing an opportunity for improvement in sepsis care among transferred patients.^68^ Future work that can improve timely sepsis diagnosis and early treatment, especially in rural sepsis patients being transferred, could be an important approach to improve sepsis outcomes.

Our subgroup analyses reveal several notable findings. Rural sepsis patients treated by APPs who had tele-ED used as part of their care had lower mortality than those that did not. This finding is relevant because APPs were more likely to use tele-ED care, and they used it earlier in the visit—suggesting that they may have been seeking more guidance in the care of these patients. The association in APP tele-ED use and guideline adherence was not statistically significant, but the point estimate of 2.45 is qualitatively different from that measured in the study overall, suggesting that care in this subgroup may actually have been higher quality. Subgroup analysis of those with tele-ED used within 1 hour was also associated with improved survival, which supports the hypothesis that early consultation is necessary for recommendations to change care. We also observed point estimates in low-volume hospitals and high-illness-severity patients that, although not statistically significant, suggest that future tele-ED comparative effectiveness studies should target high-illness-severity patients in low-volume hospitals treated by less experienced staff. The lack of statistical significance for subgroup analysis should be hypothesis-generating only, because small samples, large effect sizes, and wide confidence intervals suggest that these observations may not be replicable.

In conclusion, tele-ED consultation in rural EDs that have 24-hour access to provider-to-provider tele-ED as applied in this network was not associated with more 28-day hospital-free days or improved sepsis guideline adherence. Patients treated by APPs may represent a population in which tele-ED was more likely to be used, and the comparative effectiveness in this subgroup may be greater. Future work will focus on the role of tele-ED interventions at improving early sepsis diagnosis; heterogeneous treatment effects according to provider, hospital, and illness characteristics; and the role of network maturity in driving ongoing quality improvement activities.

## Data Availability

All deidentified data produced in the present study are available upon reasonable request to the authors.

## ACKNOWLEDGMENTS

The authors acknowledge Hannah McKay, MS, Mitchell Shaffer, BS, and Cathy Fairfield, RN, BSN for their assistance with data collection, Nathan Kramer, MPH, Paul Casella, MFA, and David Talan, MD for their editorial assistance, and Steven Q. Simpson, MD for his assistance with study design. This study was funded by the Agency for Healthcare Research and Quality (K08HS025753) and the Institute for Clinical and Translational Science at the University of Iowa through a grant from the National Center for Advancing Translational Sciences at the National Institutes of Health (UL1TR002537). Dr. Mohr is additionally supported by funding from the Rural Telehealth Research Center with funding from the Health Resources and Services Administration (UICRH29074). The funders had no role in the collection or analysis of research data. These contents are solely the responsibility of the authors and do not necessarily reflect the views of the Agency for Healthcare Research and Quality.

