## Supplemental Materials for "Outcomes Associated with Rural Emergency Department Provider-to-Provider Telehealth for Sepsis Care: A Multicenter Cohort Study"

#### TABLES

**Table S1.** Characteristics of matched and unmatched sepsis patients 2

#### FIGURES

**Figure S1.** Map of participating sites 4

**Figure S2.** Time to tele-ED consultation in rural sepsis patients 5

**Figure S3.** Distribution of sepsis cases by hospital in cohort 6

**Figure S4.** Prevalence of tele-ED use and transfer behavior 7

**Figure S5.** Distribution of propensity scores in the sample of sepsis cases 8

**Figure S6.** Standardized mean differences (SMD) of each variable in the full sample and in the propensity-matched cohort 9

**Figure S7.** Survival plot showing the proportion of tele-ED and non-tele-ED patients remaining hospitalized over time 10

**Figure S8.** Sensitivity analysis showing the relationship between tele-ED use and antibiotic appropriateness 11

**Figure S9.** Sensitivity analysis showing the relationship between tele-ED use and elements of guideline adherence 12

**Figure S10.** Distribution of sepsis cases by rural treating clinician 13

**Figure S11.** Sensitivity analysis of clinical outcomes for tele-ED patients with consultation less than 3 hours from first sepsis criteria 14

**Figure S12.** Sensitivity analyses of tele-ED use for Surviving Sepsis Campaign (SSC) guideline adherence if tele-ED was consulted within 3 hours 15

#### APPENDIX

**Appendix A.** Detailed statistical methods 16

### SUPPLEMENTAL TABLES

#### Table S1. Characteristics of matched and unmatched sepsis patients

| | Tele-ED Cases | | Non-Tele-ED Cases | |
|---|---|---|---|---|
| | All Cases (n=865) n (%) | Matched Cases (n=368) n (%) | All Cases (n=326) n (%) | Matched Cases (n=242) n (%) |
| **DEMOGRAPHICS** | | | | |
| Age | | | | |
| <30 y | 7 (0.8) | 2 (0.5) | 3 (0.9) | 1 (0.4) |
| ≥30 y and <40 y | 28 (3.2) | 15 (4.1) | 13 (4.0) | 10 (4.1) |
| ≥40 y and <50 y | 39 (4.5) | 28 (7.6) | 29 (8.9) | 20 (8.3) |
| ≥50 y and <60 y | 79 (9.1) | 42 (11.4) | 42 (12.9) | 31 (12.8) |
| ≥60 y and <70 y | 207 (23.9) | 122 (33.2) | 99 (23.6) | 68 (28.1) |
| ≥70 y and <80 y | 195 (22.5) | 81 (22.0) | 77 (23.6) | 60 (24.8) |
| ≥80 y and <90 y | 231 (26.7) | 69 (18.8) | 55 (16.9) | 44 (18.2) |
| ≥90 y | 79 (9.1) | 9 (2.5) | 8 (2.5) | 8 (3.3) |
| Female | 405 (46.8) | 154 (41.9) | 131 (40.2) | 100 (41.3) |
| Body mass index, kg/m$^2$ | | | | |
| <18.5 | 36 (4.3) | 9 (2.6) | 11 (3.1) | 6 (2.6) |
| ≥18.5 and < 25 | 227 (27.9) | 77 (22.4) | 67 (22.3) | 53 (23.4) |
| ≥25 and <30 | 220 (27.1) | 103 (29.9) | 80 (26.6) | 59 (26.0) |
| ≥30 and <35 | 142 (17.5) | 69 (20.1) | 62 (20.6) | 47 (20.7) |
| ≥35 | 190 (23.4) | 86 (25.0) | 84 (27.9) | 62 (27.3) |
| Latinx | 9 (1.1) | 8 (2.2) | 5 (1.6) | 4 (1.7) |
| Race | | | | |
| White | 773 (90.3) | 326 (90.1) | 286 (88.3) | 215 (89.2) |
| African American | 3 (0.4) | 2 (0.6) | 5 (1.5) | 3 (1.2) |
| American Indian or Alaska Native | 66 (7.7) | 29 (8.0) | 27 (8.3) | 19 (7.9) |
| Other | 14 (1.6) | 5 (1.4) | 6 (1.9) | 4 (1.7) |
| **PATIENT CHARACTERISTICS** | | | | |
| Triage systolic blood pressure, mmHg | 132 (111–151) | 128 (108 – 148) | 126 (105–146) | 126 (108 -147) |
| SOFA Score | | | | |
| 2–3 | 496 (55.0) | 159 (43.2) | 110 (33.7) | 100 (41.4) |
| 4–7 | 364 (42.1) | 196 (53.3) | 173 (53.1) | 124 (51.2) |
| ≥8 | 25 (2.9) | 13 (3.5) | 43 (13.2) | 18 (7.4) |
| Source of Infection | | | | |
| Respiratory | 502 (58.0) | 176 (47.8) | 133 (40.8) | 109 (45.0) |
| Genitourinary | 154 (17.8) | 66 (17.9) | 57 (17.5) | 47 (19.4) |
| Abdominal | 49 (5.7) | 31 (8.4) | 28 (8.6) | 18 (7.4) |
| Device-Related | 1 (0.1) | 1 (0.3) | 1 (0.3) | 1 (0.4) |
| Skin/Soft Tissue | 76 (8.8) | 44 (12.0) | 33 (10.1) | 25 (10.3) |
| Meningitis | 6 (0.7) | 3 (0.8) | 8 (2.5) | 5 (2.1) |
| Unknown | 77 (8.9) | 47 (12.8) | 66 (20.3) | 37 (15.3) |
| Surgery during hospital stay | 44 (5.1) | 17 (4.6) | 37 (11.4) | 28 (11.6) |
| Interhospital Transfer | 73 (8.4) | 60 (16.3) | 286 (87.7) | 207 (85.5) |
| Provider Type | | | | |

| | Tele-ED Cases | | Non-Tele-ED Cases | |
|---|---|---|---|---|
| | All Cases (n=865) n (%) | Matched Cases (n=368) n (%) | All Cases (n=326) n (%) | Matched Cases (n=242) n (%) |
| Physician | 781 (90.3) | 306 (83.2) | 231 (70.9) | 186 (76.9) |
| APP with physician | 59 (6.8) | 43 (11.7) | 57 (17.5) | 36 (14.9) |
| APP alone | 25 (2.9) | 19 (5.2) | 38 (11.7) | 20 (8.3) |
| Arrive by Ambulance | 416 (49.2) | 175 (47.6) | 167 (51.9) | 116 (47.9) |
| **COMORBIDITIES** | | | | |
| COPD | 292 (33.8) | 104 (28.3) | 86 (26.4) | 70 (28.9) |
| Cirrhosis | 18 (2.1) | 12 (3.3) | 13 (4.0) | 6 (2.5) |
| Solid organ transplant | 8 (0.9) | 5 (1.4) | 4 (1.2) | 2 (0.8) |
| Cancer | 266 (30.8) | 100 (27.2) | 88 (27.0) | 70 (28.9) |
| Diabetes | 314 (36.3) | 136 (37.0) | 126 (38.7) | 97 (40.1) |
| Chronic Dialysis | 21 (2.4) | 15 (4.1) | 24 (7.4) | 15 (6.2) |
| Asthma | 103 (11.9) | 41 (11.1) | 40 (12.3) | 30 (12.4) |
| Hypertension | 631 (73.0) | 253 (68.8) | 225 (69.0) | 165 (68.2) |

### SUPPLEMENTAL FIGURES

**Supplemental Figure S1. Map of participating sites.**

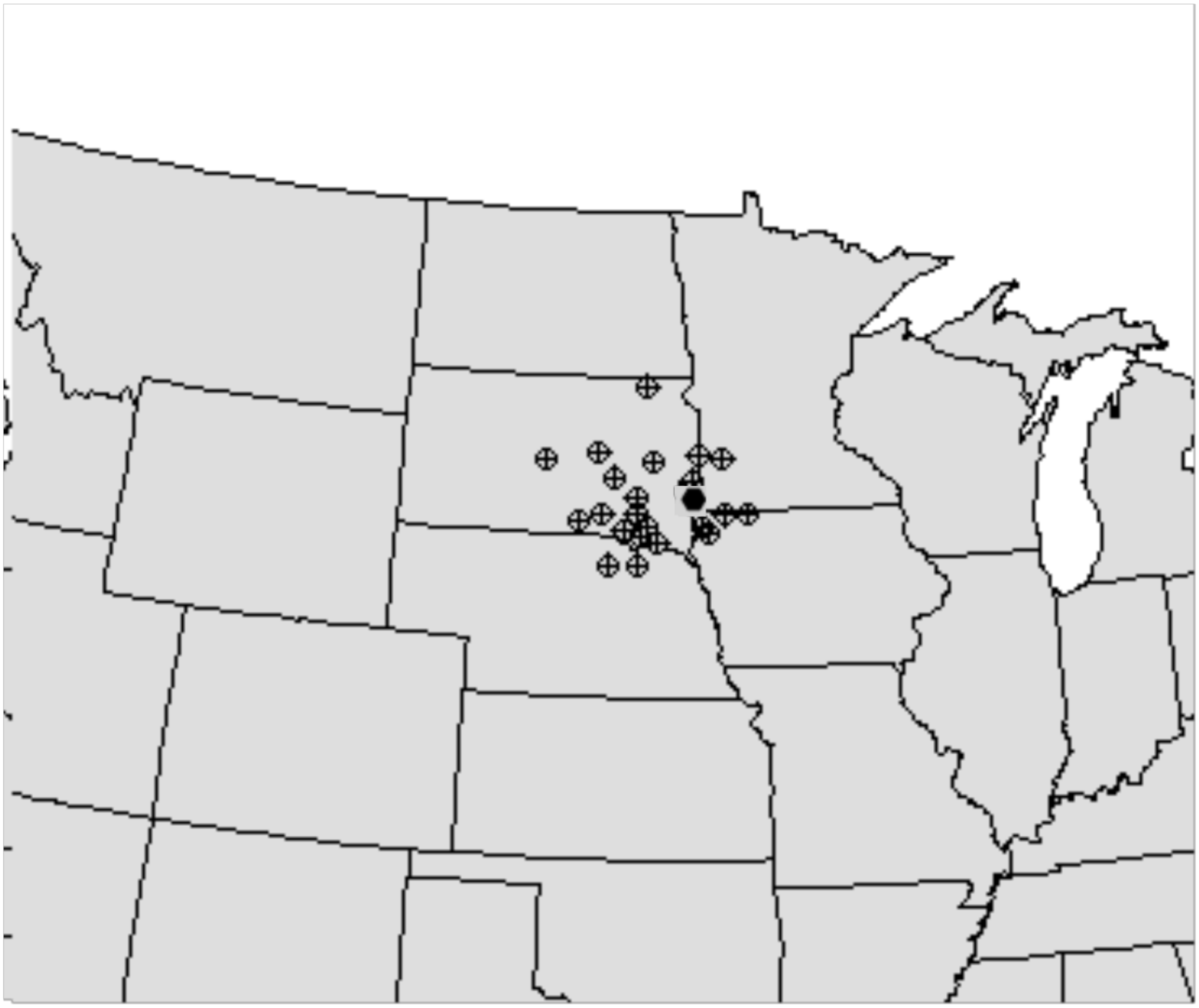

**Supplemental Figure S2. Time to tele-ED consultation in rural sepsis patients.** Each bar in this histogram represents the percentage of all tele-ED patients for whom tele-ED was activated within a given time from first documented sepsis criteria in the ED. Seventy-one percent of tele-ED consultations occurred after 60 minutes, and 18% occurred after 180 minutes.

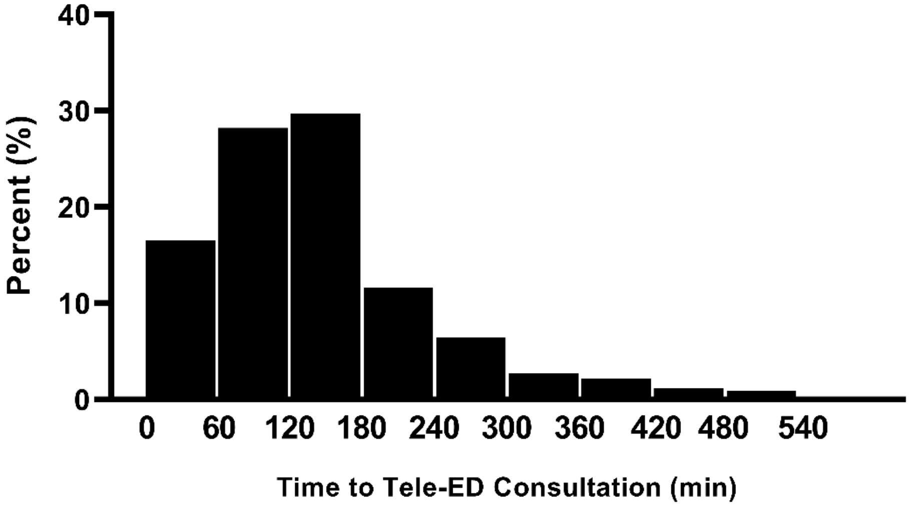

**Supplemental Figure S3. Distribution of sepsis cases by hospital in cohort.** The black bars represent the number of sepsis cases for which tele-ED was used, and the gray bars represent the number of sepsis cases for which tele-ED was not used in each hospital. Each bar along the horizontal axis represents one hospital in the cohort.

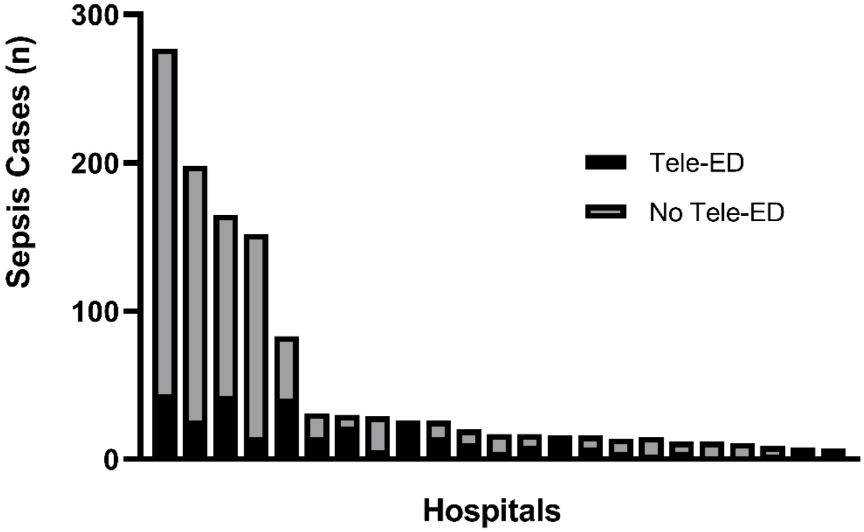

**Supplemental Figure S4. Prevalence of tele-ED use and transfer behavior.** In this graph, each dot represents one participating hospital. The horizontal axis shows the prevalence of inter-hospital transfer, and the vertical axis shows the corresponding prevalence of tele-ED use among sepsis patients. The size of the dot represents the number of sepsis cases treated in the participating emergency department. This graph represents that hospitals that used tele-ED most frequently were smaller hospitals that were also more likely to transfer sepsis patients for care.

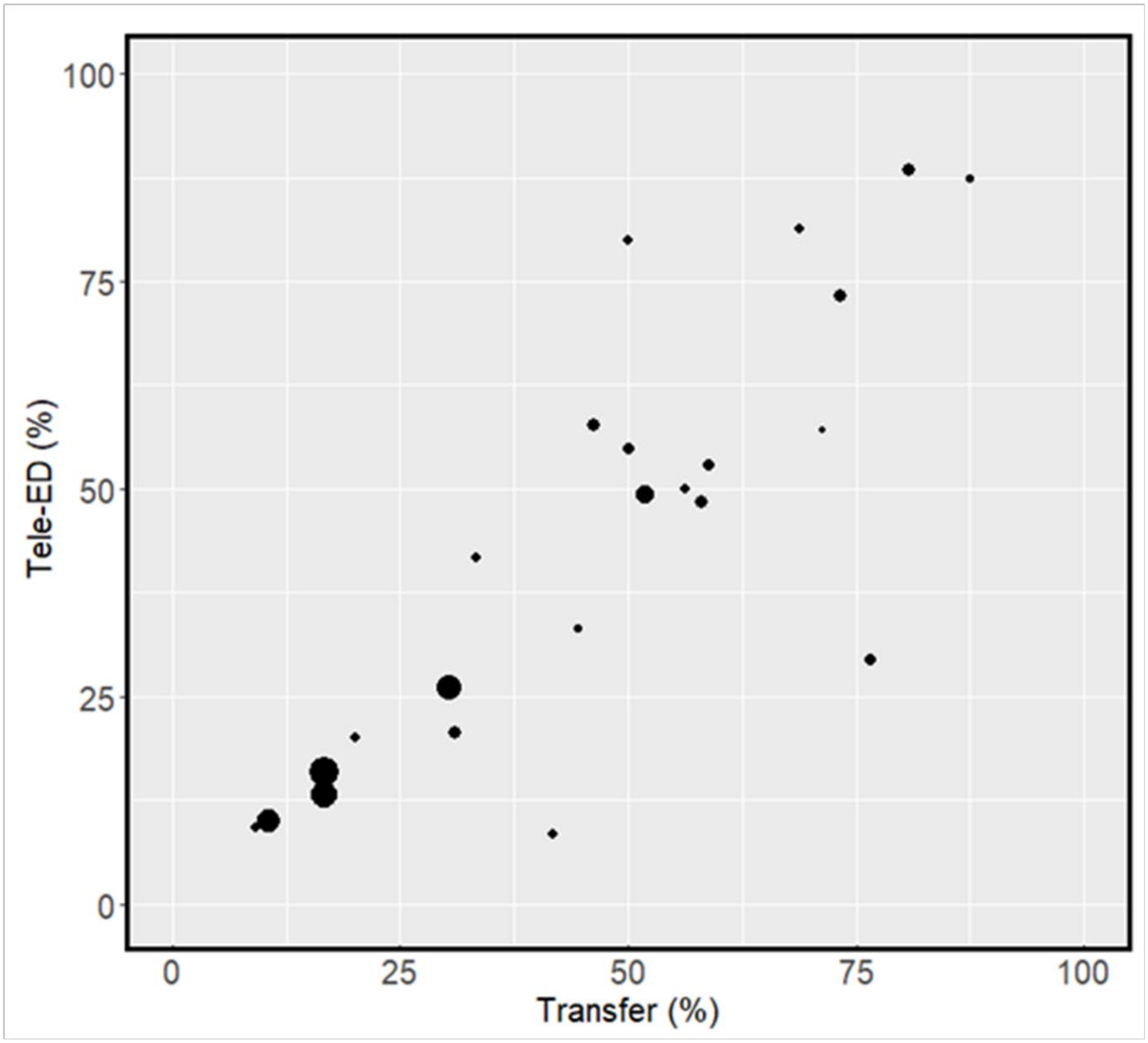

**Supplemental Figure S5. Distribution of propensity scores in the sample of sepsis cases.** The panel on the left shows the distribution of propensity scores for the entire sample, and the panel on the right shows the distribution of scores among the matched cohort. The dark gray distribution is tele-ED cases, and the light gray distribution is the non-tele-ED cases. This graph represents data from the first imputation only, but the distributions were similar for other imputations. *Tele-ED, provider-to-provider emergency department-based telemedicine consultation*

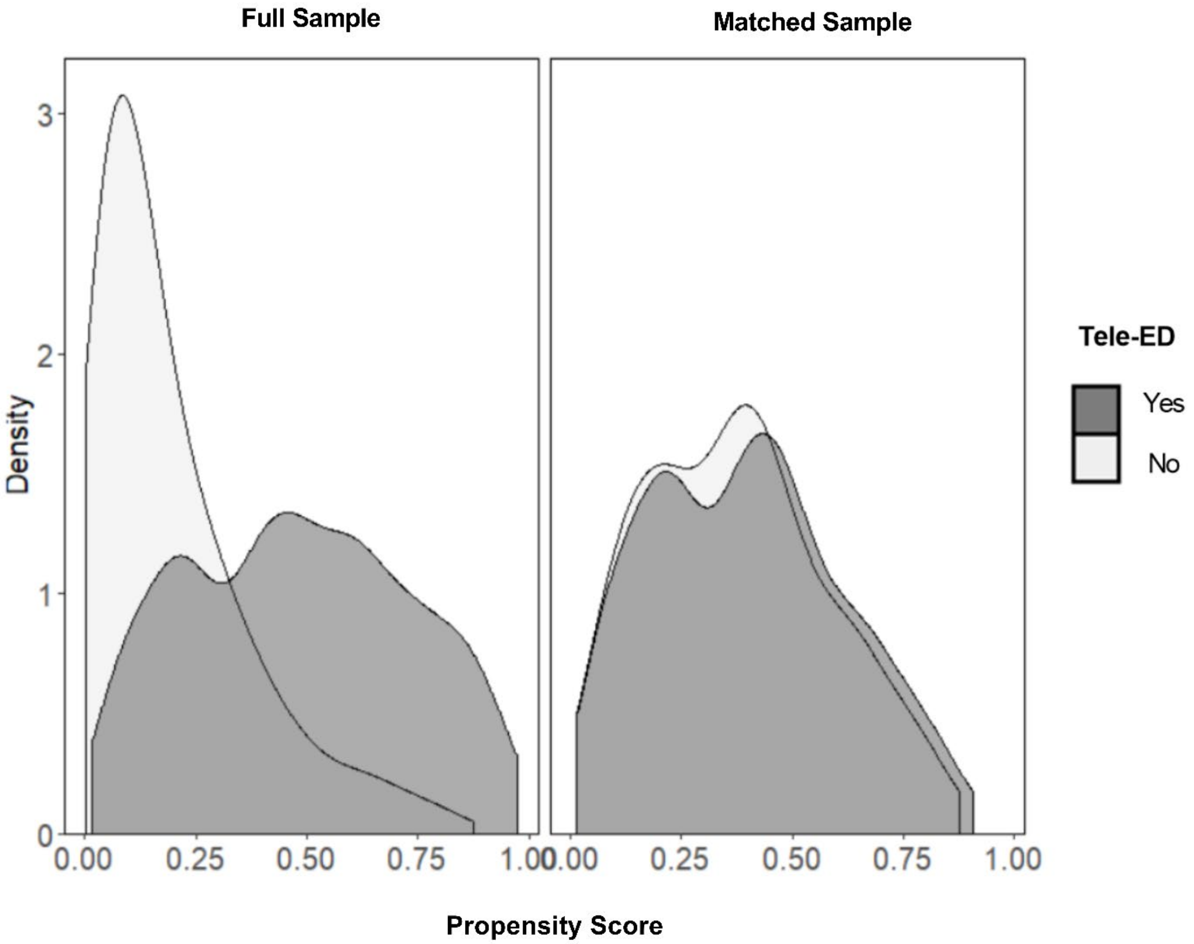

**Supplemental Figure S6. Standardized mean differences (SMD) of each variable in the full sample and in the propensity-matched cohort.** SMD<0.1 (region bounded by the dotted lines) generally represents good matching. This graph represents data from the first imputation only, but the distributions were similar for other imputations. *BMI, body mass index; ED, emergency department; SOFA, sequential organ failure assessment; CNS, central nervous system; COPD, chronic obstructive pulmonary disease; ICU, intensive care unit*

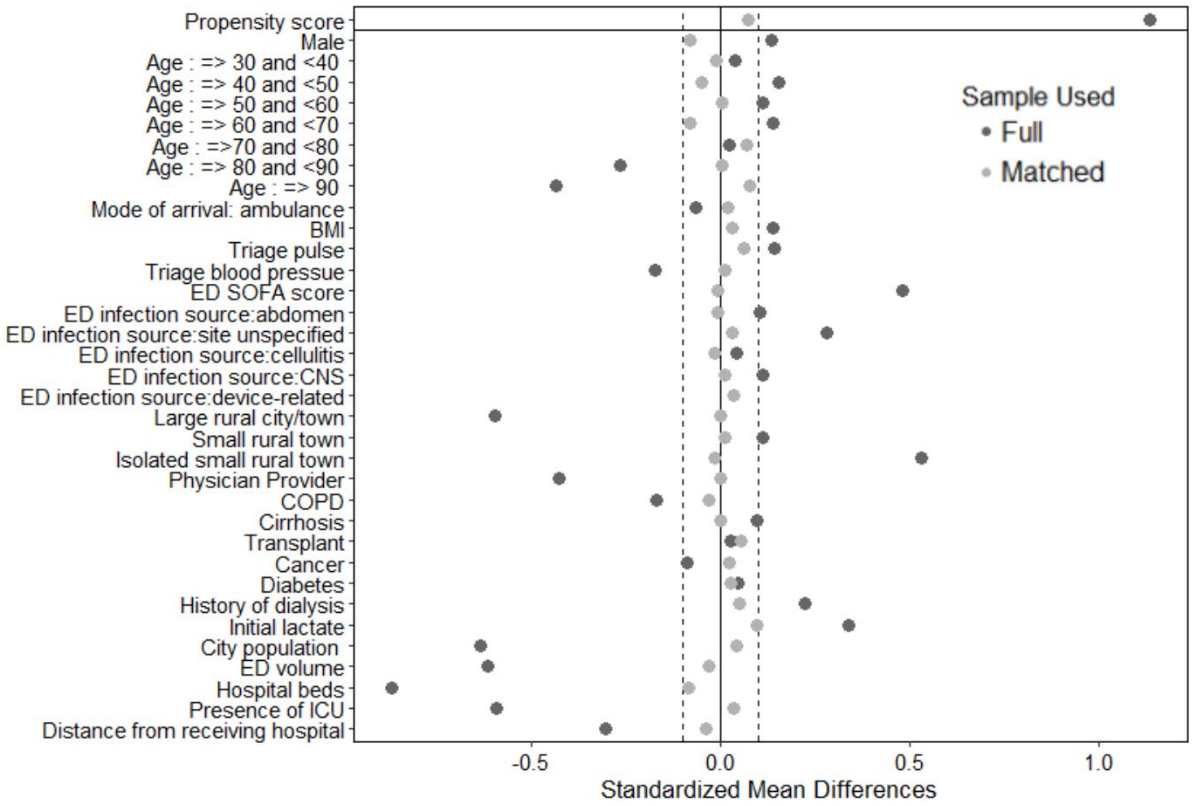

**Supplemental Figure S7. Survival plot showing the proportion of tele-ED and non-tele-ED patients remaining hospitalized over time.** This figure shows unadjusted data representing hospital length-of-stay. Sepsis patients who died are censored, and they are indicated with a vertical hash mark. Hospitalization for this plot includes length-of-stay at all hospitals (including those to which patients were transferred). All tele-ED consultations (for the tele-ED group) were provided on Day 0. *d, days; tele-ED, provider-to-provider telemedicine consultation in rural emergency department*

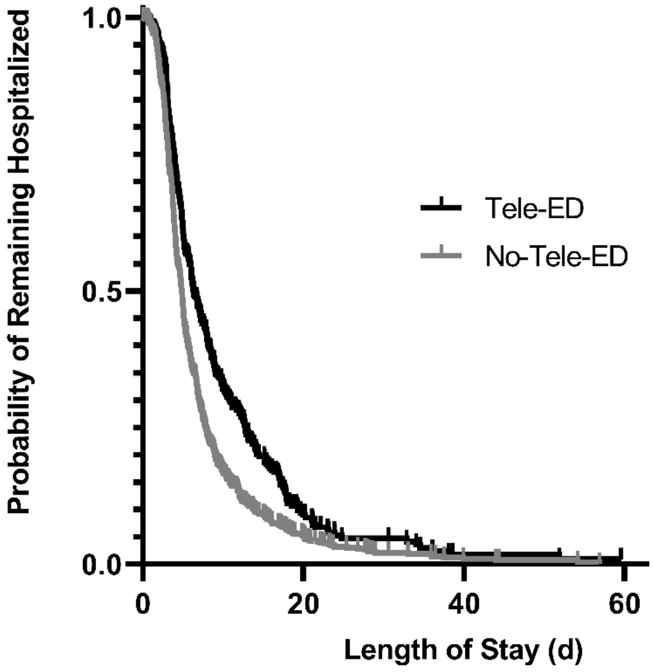

**Supplemental Figure S8. Sensitivity analysis showing the relationship between tele-ED use and antibiotic appropriateness.** The bar chart on the left shows the proportion of total patients who were treated with appropriate antibiotics, stratified by tele-ED use. The forest plot on the right side shows the relationship between tele-ED use and antibiotic appropriateness, using each of the 2 definitions for antibiotic appropriateness. Grey circles represent unadjusted associations (measured in the full unmatched data set), and black squares represent the adjusted models in the propensity-matched cohort.

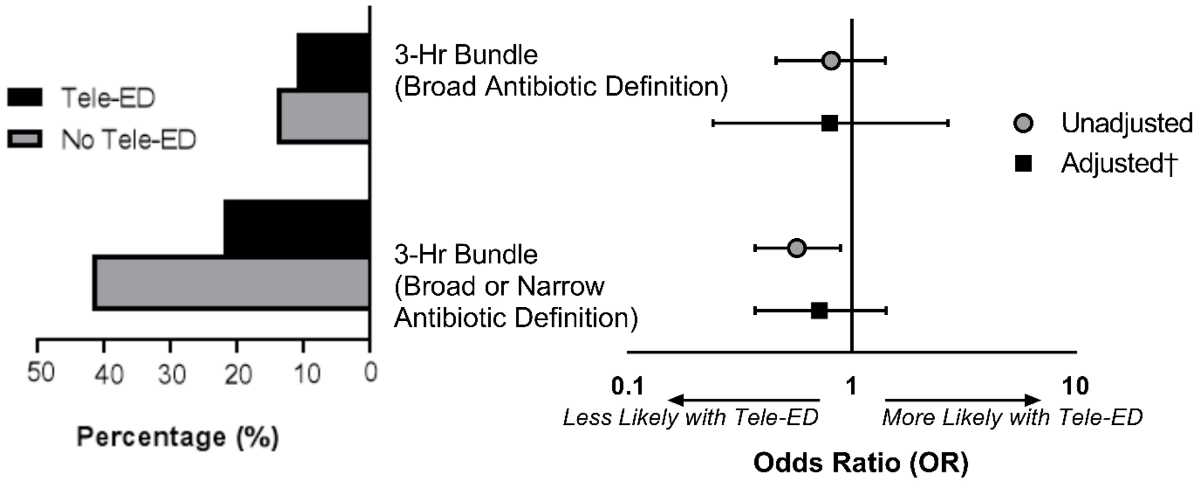


†The adjusted model included the following covariates in the propensity-matched cohort: ED annual volume, index hospital inpatient beds, age, and interhospital transfer.

**Supplemental Figure S9. Sensitivity analysis showing the relationship between tele-ED use and elements of guideline adherence.** In this sensitivity analysis, we measured the relationship between tele-ED use and completion of bundle elements by 6 hours only. Grey circles represent unadjusted associations (measured in the full unmatched data set), and black squares represent the adjusted models in the propensity-matched cohort. The last row shows complete guideline adherence by 6 hours (late).

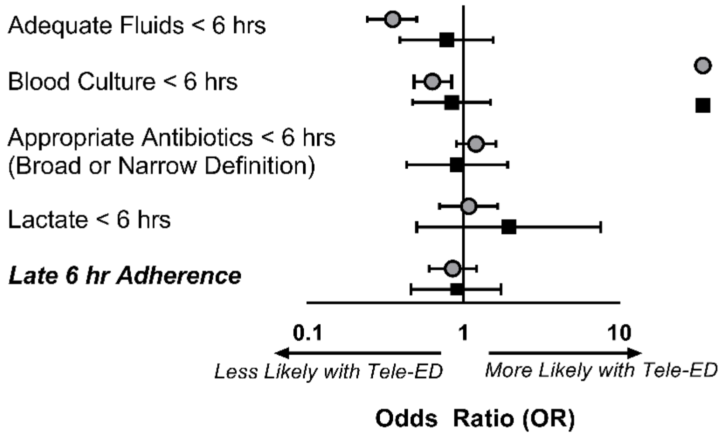


†The adjusted model included the following covariates in the propensity-matched cohort: ED annual volume, index hospital inpatient beds, age, and interhospital transfer.

**Supplemental Figure S10. Distribution of sepsis cases by rural treating clinician.** In this graph, black bars represent the number of patients in each provider classification that used tele-ED, and gray bars represent the number of cases that did not use tele-ED. The percentages to the right of the bars represent the proportion of cases seen by that provider type that had tele-ED consulted. *APP, advanced practice provider; n, count*

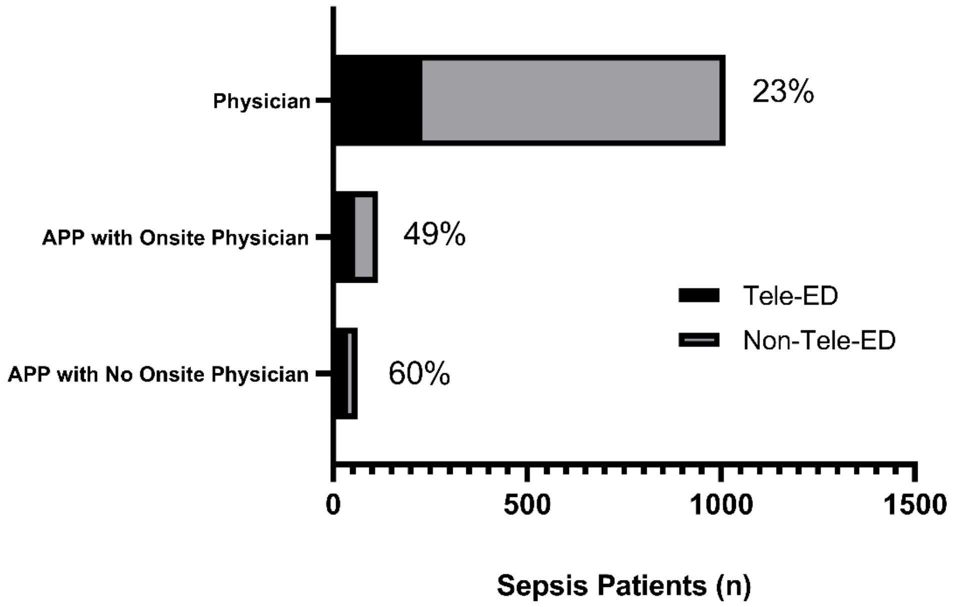

**Supplemental Figure S11. Sensitivity analysis of clinical outcomes for tele-ED patients with consultation less than 3 hours from first sepsis criteria.** The forest plots show the association between tele-ED use and clinical outcomes. Grey circles represent unadjusted associations (measured in the full unmatched data set), and black squares represent the adjusted models in the propensity-matched cohort. **A.** Count or continuous outcomes. **B.** Dichotomous outcomes.
*SOFA, sequential organ-failure assessment score*

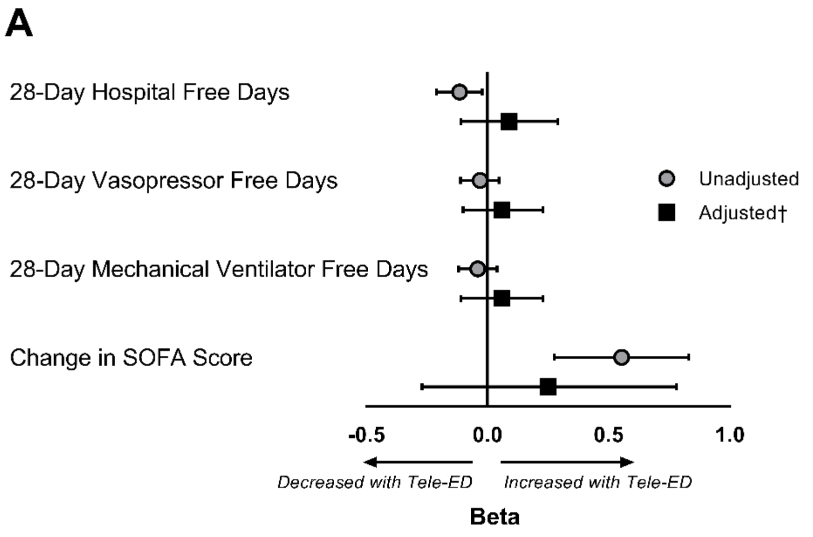


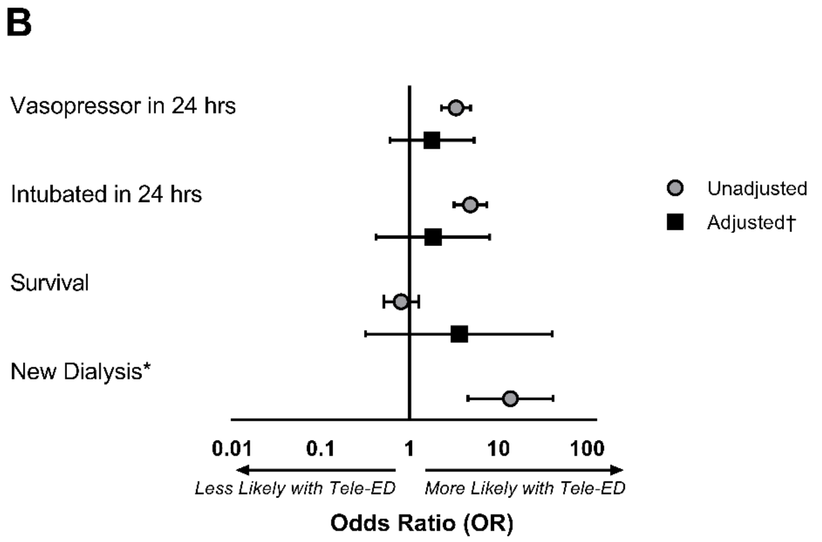


*The adjusted model for new dialysis did not converge because there were very few outcomes in the data set.
†The adjusted model included the following covariates in the propensity-matched cohort: ED annual volume, index hospital inpatient beds, age, and interhospital transfer.

**Supplemental Figure S12. Sensitivity analyses of tele-ED use for Surviving Sepsis Campaign (SSC) guideline adherence if tele-ED was consulted within 3 hours.** The bar chart on the left side of the figure shows the proportion of tele-ED and non-tele-ED cases in the entire data set that had each element of the SSC bundles completed. On the right side, the forest plot shows the unadjusted association between tele-ED care and guideline adherence (in the matched sample only) and the adjusted association was calculated with a logistic regression model, adjusting for ED annual volume, index hospital inpatient beds, age, and interhospital transfer.

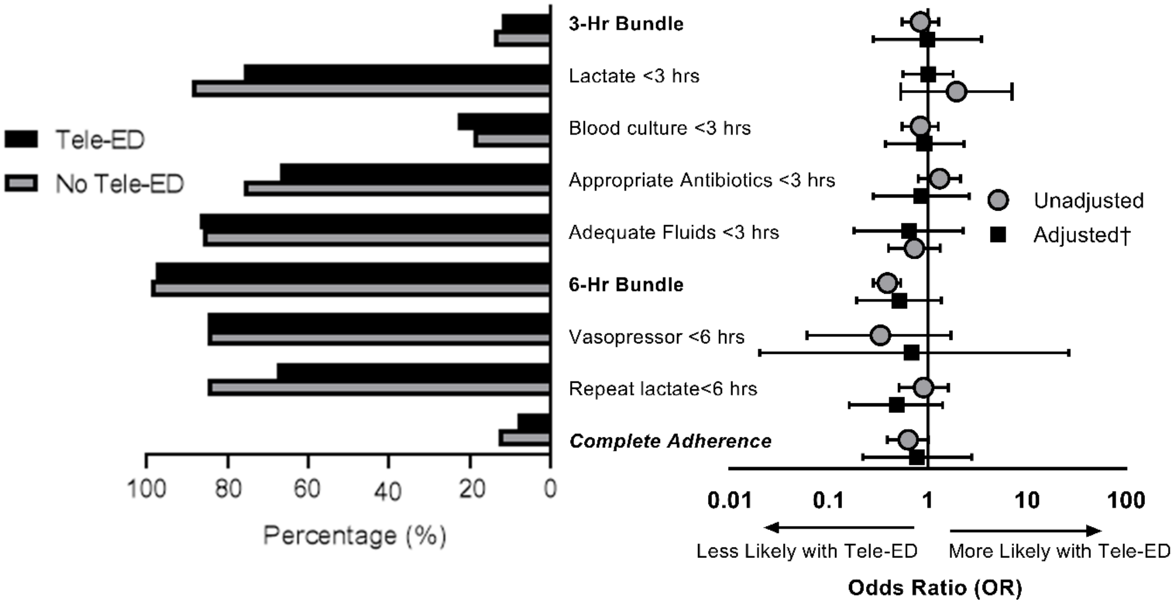


†The adjusted model included the following covariates in the propensity-matched cohort: ED annual volume, index hospital inpatient beds, age, and interhospital transfer.

### Supplemental Appendix A. Detailed Statistical Methods

Propensity Score Matching. We used a propensity-matched cohort design to adjust for differences in potentially confounding covariates. We included the following variables in our propensity score: age (by decade), sex, body mass index (BMI, 5 categories), past medical history (COPD, cirrhosis, solid organ transplant, cancer, diabetes, chronic dialysis), mode of arrival (private vehicle vs. ambulance), triage pulse (4 categories), triage systolic blood pressure, ED SOFA score (3 categories), initial lactate (3 categories), suspected source of infection (respiratory/pneumonia , genitourinary/urinary tract infection, abdominal, device-related, endocarditis, wound/soft tissue/cellulitis, central nervous system/meningitis, site unspecified), and ED provider type (physician, APP with onsite physician, APP without onsite physician).

Initially, we intended to identify exact matches within hospital to account for baseline differences in care within a hospital, but insufficient matches existed. Next, we attempted to include the hospital identifier in the propensity score, but match efficiency was still poor. Finally, we developed a model including hospital characteristics instead of the hospital identifier, and the area under the curve (AUC) of the regression model approximated a similar model including the hospital identifier (0.735 vs. 0.759). Because of that, we added the following hospital-level variables to the propensity score: hospital rurality (measured with the rural-urban commuting areas[52]), city population, ED annual volume, hospital inpatient beds, presence of an ICU in the index hospital, and distance from the most likely receiving hospital.

All continuous variables except for blood pressure, ED annual volume, index hospital inpatient beds, and distance from the index hospital to the most likely receiving hospital were categorized for inclusion in the propensity score because they did not behave linearly in predicting the outcome. After development of our model using categorized continuous variables, we compared the fit of a model with all continuous variables included linearly with our model with categorized covariates, and we found that the model with categorized covariates fit better (AUC 0.832 vs. 0.817).

Because significantly more tele-ED non-exposed cases were available in the analytic data set, we matched tele-ED cases in a variable ratio (nominal 1:2 matching ratio when suitable controls were available) with non-exposed controls using an optimal matching algorithm to minimize overall Mahalanobis distance. To ensure that the optimal algorithm did not match cases that were dissimilar, we included a maximum caliper width of 0.15, which led to some broken matches (less than 1:2 allocation in our final sample). We measured standardized mean differences in individual variables and in the overall propensity score to ensure balance, and we also manually reviewed individual matches to confirm that matched cases were qualitatively similar.

Missing Data. We conducted a complete case analysis on outcomes. We did permit cases with missing covariates, however, to be included in the analysis. We used multiple imputation to generate independent data sets for matching and regression modeling, then we pooled the results to determine effect estimates at the end of analysis. For this analysis, we used 5 imputed data sets, calculated using the Von Hippel method.[69] For transparent reporting, we report only one imputation for matching diagnostics, but the final estimates reflect the pooled result.

Imbalance in Inter-Hospital Transfer. During analysis, we realized that inter-hospital transfer is strongly related to tele-ED use. Initially, we planned to leave this variable out of our matching algorithm because transfer occurs *after* tele-ED care was delivered, but after talking with experts who work inside the tele-ED network, we realized that tele-ED is often consulted for assistance with the transfer process (e.g., reverse causality). We also realized in our early matched cohorts that inter-hospital transfer exhibited significant imbalance, and we thought the magnitude of the imbalance made a causal relationship between tele-ED and transfer unlikely. Because of these considerations, we decided to include inter-hospital transfer in the statistical adjustment for the analysis because we assumed that it reflected information available to the rural clinician, but not captured in other covariates in our model. We attempted to include that variable in the propensity score, but the distribution of scores was strongly bimodal, match efficiency was low, and balance on all the non-transfer variables suffered significantly. We then tried to force exact matches on inter-hospital transfer, but match efficiency remained low and balance on non-transfer variables remained poor. Finally, we elected to include inter-hospital transfer as a right-sided predictor variable in our regression analysis for each of our outcome models.

Regression Modeling. We modeled each of our outcomes using generalized linear models (GLM) within the propensity-matched cohort clustered on propensity-matched pair identifier. Any variables with standardized mean difference >0.1 in our matched cohorts were included as right-sided adjustment variables, and inter-hospital transfer was forced into the model. We used a negative binomial distribution with log link for our count outcomes (e.g., 28-day hospital-free days) because our data were over-dispersed, and we used a logit model for dichotomous outcomes. Each primary and secondary outcome had a similar model constructed (using the same covariates).

Subgroup Analysis. For the predefined subgroup analysis of patients treated by APPs, we decided *a priori* to use traditional GLM regression techniques only (without propensity matching) because we anticipated low counts. This analysis was conducted with the primary outcome of 28-day in-hospital mortality and secondary outcome of complete guideline adherence). We intended to use the same technique for cases in low-volume hospitals (with case counts <100 over study period), cases with high illness severity (SOFA score >6), and for those tele-ED cases with hub-recognized sepsis if the proportion of total ED cases was low.

For the subgroup of tele-ED cases consulted within 3 hours of sepsis diagnosis, we replicated our original propensity-matched analysis, but excluding any tele-ED cases consulted after 3 hours. We matched remaining cases with controls and built similar regression models.
